# Interleukin-2 immunotherapy reveals human regulatory T cell subsets with distinct functional and gatekeeper features

**DOI:** 10.1101/2022.11.15.22282201

**Authors:** Miro E. Raeber, Dominic Caspar, Yves Zurbuchen, Nannan Guo, Jonas Schmid, Jan Michler, Urs C. Steiner, Andreas E. Moor, Frits Koning, Onur Boyman

**Affiliations:** Department of Immunology, University Hospital Zurich, Zurich, Switzerland; Department of Immunology, Leiden University Medical Center, Leiden, Netherlands; Department of Biosystems Science and Engineering, ETH Zurich, Basel, Switzerland; Faculty of Medicine and Faculty of Science, University of Zurich, Zurich, Switzerland

## Abstract

Due to its stimulatory potential for immunomodulatory CD4^+^ regulatory T (Treg) cells, low-dose interleukin-2 (IL-2) immunotherapy has recently gained considerable attention for treatment of various autoimmune diseases. Although early-stage clinical trials have correlated expansion of circulating Treg cells with clinical response to IL-2 treatment, detailed mechanistic data on responding Treg cell subsets are lacking. In this investigator-initiated phase-2 clinical trial of low-dose IL-2 immunotherapy in systemic lupus erythematosus (SLE) patients, we performed an in-depth study of circulating and cutaneous Treg cell subsets by imaging mass cytometry, high-parameter spectral flow cytometry, bulk and single-cell RNA sequencing with cellular indexing, and targeted serum proteomics. Low-dose IL-2 stimulated circulating Treg cells with skin-homing properties that appeared in the skin of SLE patients in close interaction with endothelial cells, suggestive of a gatekeeper function. Analysis of surface proteins and transcriptomes detected different IL-2-driven Treg cell programs, including highly proliferative CD38^+^ HLA-DR^+^, activated gut-homing CD38^+^, and skin-homing HLA-DR^+^ Treg cells. These data identify distinct and functionally characteristic Treg cell subsets in human blood and skin, including the Treg cell subsets most responsive to IL-2 immunotherapy, thus providing unprecedented insight into Treg cell biology during IL-2 treatment.

## INTRODUCTION

Although interleukin-2 (IL-2) was tried at high doses first in immunotherapy of cancer patients, the discovery of IL-2-dependent CD4^+^ regulatory T (Treg) cells has spurred efforts to use low-dose IL-2 treatment in various autoimmune diseases^1-4^. These apparently contrasting effects are mechanistically explained by the two main configurations of the IL-2 receptor (IL-2R). A dimeric, intermediate-affinity IL-2R, consisting of IL-2Rβ (CD122) and IL-2Rγ (CD132), is mainly expressed on effector T (Teff) cells and natural killer (NK) cells, whereas a trimeric, high-affinity IL-2R also contains IL-2Rα (CD25) and is mainly expressed on CD4^+^ forkhead box P3 (FOXP3)^+^ Treg cells and innate lymphoid cells^5-7^. Due to this distinct distribution of IL-2R, low doses of IL-2 preferentially expand Treg cells, while high doses additionally stimulate effector T and NK cells^3,7^. Several clinical trials testing low-dose IL-2 in different autoimmune diseases have confirmed preferential expansion of Treg cells in peripheral blood; however, they did not provide an in-depth characterization of IL-2-induced phenotypical and functional changes of circulating and tissue Treg cells, compared to other lymphocyte subsets^8-11^.

Systemic lupus erythematosus (SLE) is a multi-system autoimmune disease, preferentially affecting women of childbearing age. SLE can damage various organs, including the skin, joints, kidneys, and central nervous system. Secondary autoimmune hemolytic anemia and immune thrombocytopenia are also frequently observed. Due to the presence of pathognomonic autoantibodies, including antinuclear antibodies (ANA), anti-double-stranded DNA (dsDNA) antibodies, and anti-Smith antibodies, SLE was considered a prototypical B cell-mediated disease^12,13^. However, more detailed immune phenotyping in recent years and the limited success of initial B cell-depleting biologics have revised this perspective, revealing the involvement of various immune cells in the pathogenesis of SLE^14-16^. In particular, the discovery of a functional deficit of Treg cells in SLE defined by lower CD25 expression and a deficiency in homeostatic proliferation compared to CD4^+^ conventional T (Tcon) cells, both of which correlated with disease activity, prompted the first clinical trial to test low-dose IL-2 for Treg cell restoration in SLE^8,9^. Additional trials followed, however, two recent phase-2 randomized controlled trials did not meet their primary endpoint of clinical efficacy^9,17-20^. A reason for these outcomes could be the lack of data identifying precise Treg cell subsets, including those that proliferate most upon IL-2 immunotherapy and migrate to target organs in SLE, thus allowing specific patient stratification^2^.

By exploiting recent advances in imaging mass cytometry, spectral flow cytometry, single-cell RNA sequencing, and targeted serum proteomics, this study provides a comprehensive characterization of IL-2-induced immunological changes on a single-cell level in peripheral blood and skin of SLE patients, revealing multiple IL-2-driven Treg cell programs with distinct migrational and functional properties.

## RESULTS

### IL-2 therapy improves SLE disease activity and promotes skin homing of Treg cells

Within the investigator-initiated clinical trial Charact-IL-2 (ClinicalTrials.gov Identifier: NCT03312335), we treated 12 female SLE patients with recombinant low-dose IL-2, with patients receiving 4 cycles of 5 daily injections of 1.5 million international units aldesleukin over 9 weeks. Skin punch biopsies and blood samples were collected and analyzed using imaging mass cytometry, high-parameter flow cytometry, bulk and single-cell RNA sequencing with cellular indexing, and targeted serum proteomics (**Fig. 1a**). Cyclic administration of IL-2 immunotherapy induced a pronounced expansion of CD3^+^ CD4^+^ CD127^lo^ FOXP3^+^ CD25^+^ Treg cells, with the Charact-IL-2 trial reaching its predefined primary endpoint of significant Treg cell expansion between day 0 (baseline) and day 68 (endpoint) after treatment initiation (**Fig. 1b**). IL-2 immunotherapy both reduced disease activity, as measured with the validated clinical scores SELENA-SLEDAI, BILAG-2004, and Physician Global Assessment, and improved SLE-associated serum biomarkers, thus increasing complement component 3c (C3c), but not C4, and decreasing anti-dsDNA antibodies (**Fig. 1c**,**d**). Furthermore, IL-2 treatment allowed for a reduction of concomitant daily prednisone doses (**Fig. 1e**). Clinical adverse events were generally mild and transient, with injection site reactions, flu-like symptoms, and arthralgia reported most frequently.

**Fig. 1.**
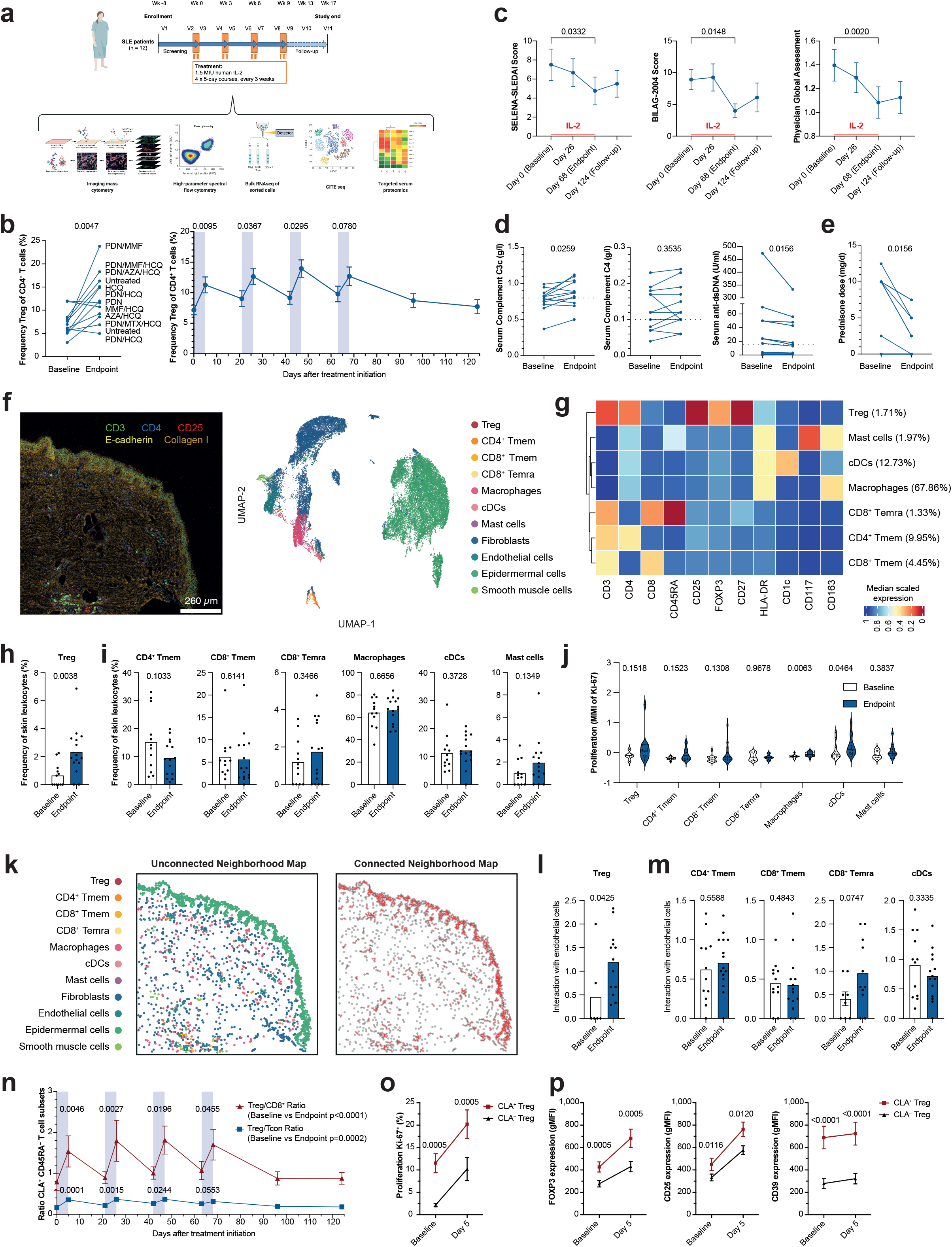
IL-2 therapy improves SLE disease activity and promotes skin homing of Treg cells. **a**, Clinical trial design with patients receiving four cycles (orange boxes) of five daily injections of 1.5 million international units of recombinant IL-2. **b**, Regulatory T (Treg) cell frequencies in blood before (day 0; baseline) and after completion (day 68; endpoint) of IL-2 immunotherapy (*n* = 12). **c**, SELENA-SLEDAI, BILAG-2004, and Physician Global Assessment scores during IL-2 treatment (*n* = 12). **d**, Changes in serological markers associated with disease activity. Dashed lines represent lower limit of normal values (*n* = 12). **e**, Daily prednisone dose (*n* = 12). **f**, Representative image and Phenograph clustering of imaging mass cytometry-analyzed paired skin punch biopsies from 4 patients taken at days 0 and 68. **g**, Heatmap depicting marker expression of immune cells identified by clustering in f. Numbers in brackets refer to frequencies of skin leukocytes. **h**,**i**, Frequency of Treg cells (h) and other leukocyte subsets (i) of total skin leukocytes (*n* = 12-15). **j**, Proliferation of skin leukocyte subsets measured by mean metal intensity (MMI) of Ki-67 (*n* = 6-15). **k**, Representative images of unconnected and connected neighborhood maps of skin leukocytes. **l**,**m**, Interaction of Treg (l) and other leukocyte subsets (m) with endothelial cells in the skin (*n* = 6-15). **n**, Increased ratios of CD3^+^ T cell frequencies of skin-homing CLA^+^ CD45RA^−^ Treg over CLA^+^ CD45RA^−^ CD8^+^ and CLA^+^ CD45RA^−^ CD4^+^ FOXP3^−^ conventional T (Tcon) cells in blood during IL-2 immunotherapy (*n* = 11-12). **o**,**p**, Proliferation (Ki-67) (o) and expression of FOXP3, CD25, and CD39 (p) of CLA^+^ compared to CLA^−^ Treg cells before and after IL-2 treatment (*n* = 11-12). Shown are mean values with error bars representing the standard error of the mean (b,c,h,I,l,m,n,o,p) or median with quartiles and minimal to maximal extension (j). After normality testing, p-values were calculated with paired t-test (b,c,d,p), Wilcoxon test (c,d,e,o,p), Mann-Whitney test (h,i,j,l,m), unpaired t-test (i,m) and Friedman test with uncorrected Dunn’s test (n).

To investigate IL-2-induced effects in tissular immune cells, we applied a 38-marker imaging mass cytometry panel to characterize immune cells in skin samples obtained before (baseline) and 9 weeks after IL-2 treatment (day 68, primary endpoint). Segmentation and clustering allowed clear distinction of immune cell subsets from non-immune cells, including fibroblasts, endothelial cells, epidermal cells, and smooth muscle cells (**Fig. 1f**). Marker expression on clustered CD4^+^ memory T (Tmem), Treg, CD8^+^ Tmem, and CD8^+^ CD45RA^+^ effector memory T (Temra), as well as on macrophages, conventional dendritic cells (cDC), and mast cells confirmed accurate annotation (**Fig. 1g**). Evaluating frequencies of these immune cell subsets in skin before and after IL-2 immunotherapy revealed a significant increase of Treg cells (**Fig. 1h**), with only minimal effects on other subsets (**Fig. 1i**). Notably, expression of the proliferation marker Ki-67 did not change significantly in skin-infiltrating Treg cells following IL-2 immunotherapy compared to baseline (**Fig. 1j**), thus lending support for improved skin homing of circulating Treg cells, rather than local expansion of skin-resident Treg cells. Interestingly, as previously reported for cDCs in mice^21^, cutaneous cDCs and macrophages increased Ki-67 abundance after IL-2 immunotherapy (**Fig. 1j**). We extended our analysis by modeling cell-cell interactions, applying a connected neighborhood map (**Fig. 1k**). This revealed increased interaction of Treg with endothelial cells after IL-2 immunotherapy (**Fig. 1l**), suggesting recent tissue infiltration through blood vessels, which further supported migration, and strategic positioning of Treg cells as gatekeepers. This effect was not observed for CD4^+^ Tmem, CD8^+^ Tmem, CD8^+^ Temra, and cDCs (**Fig. 1m**).

With the results suggesting IL-2 promoted skin infiltration of circulating Treg cells, we analyzed skin-homing markers on circulating Treg cells. We observed selective expansion of cutaneous lymphocyte-associated antigen (CLA)-expressing CD45RA^−^ Treg cells compared to CLA^+^ CD45RA^−^ CD8^+^ T and CLA^+^ CD45RA^−^ CD4^+^ FOXP3^−^ Tcon cells in the blood (**Fig. 1n**). Comparison of CLA^+^ and CLA^−^ Treg cells revealed higher functional activation of CLA^+^ Treg cells, including increased proliferation as evidenced by Ki-67 and increased suppressive properties as measured by FOXP3, CD25, and CD39 expression (**Fig. 1o**,**p**).

In summary, these results show clinical response of SLE patients to low-dose IL-2 immunotherapy correlates with preferential expansion of circulating Treg cells with skin-homing properties and increased frequencies of cutaneous Treg cells in close contact to endothelial cells in the skin.

### IL-2 immunotherapy induces modulation of immune cells

To conduct a comprehensive immune phenotyping we applied a high-dimensional spectral flow cytometry panel that allowed for the identification of all major immune cell subsets by manual gating. As circulating immune cells constitute only a minority of all immune cells and isolated analysis of peripheral cell frequencies is sensitive to bias due to treatment-induced migration to lymphoid organs and tissues, we extended our analysis to include cell proliferation measured by Ki-67 expression (**Fig. 2a**). Treg cells and CD56^bright^ NK cells showed a pronounced increase in frequency and proliferation, whereas CD4^+^ Tcon, CD8^+^ T, and CD56^+^ T cells showed only moderate signs of proliferation and did not change in terms of frequency within peripheral blood mononuclear cells (PBMC). CD56^dim^ NK cells decreased in frequency but showed very pronounced proliferation. A similar trend was observed for circulating B cells, where frequencies decreased while proliferation tended to increase gradually (**Fig. 2a**). Surprisingly, ranking proliferation after the first cycle of IL-2 immunotherapy (day 0 vs. day 5) according to level of significance revealed that CD56^bright^ and CD56^dim^ NK cells were the most IL-2-sensitive populations, directly followed by Treg cells (**Fig. 2b**). Comparing early effects after initiation of low-dose IL-2 immunotherapy (day 0 vs. day 5) revealed IL-2-induced proliferation of all investigated CD4^+^ and CD8^+^ T cell subsets, except for naïve CD4^+^ T, follicular helper T (Tfh), and recent thymic emigrant (RTE) CD4^+^ Tcon (RTE CD4^+^ Tcon) cells (**Fig. 2b**). This effect was less pronounced during the last treatment cycle of IL-2 immunotherapy (day 63 vs. day 68) (**Fig. 2c**), which could be explained by progressive expansion of Treg cells suppressing the stimulation of these effector T cell populations at late time points.

**Fig. 2.**
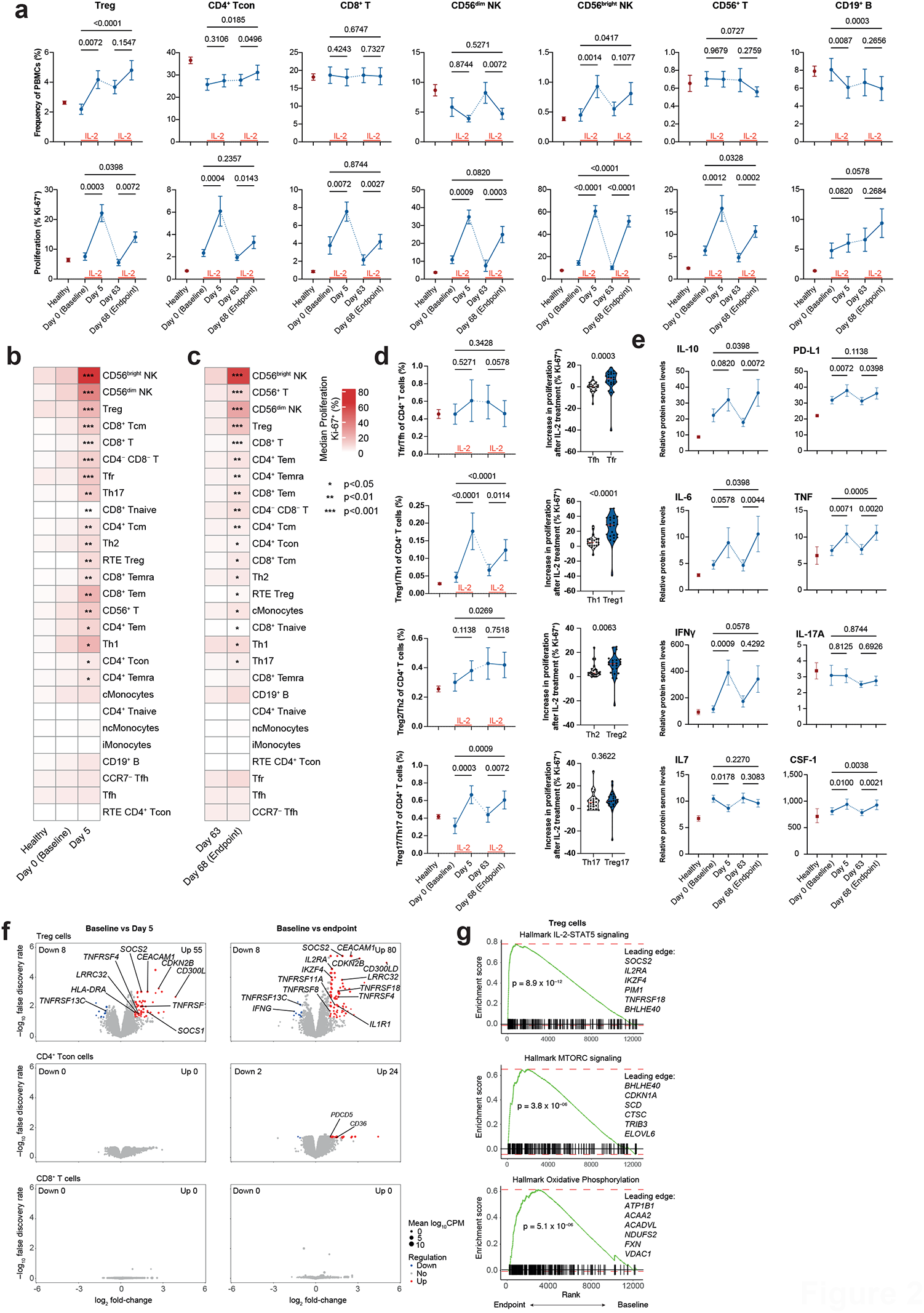
IL-2 immunotherapy induces modulation of immune cells. **a**, Frequencies of total peripheral blood mononuclear cells (PBMCs) and proliferation (% Ki-67) of selected lymphocyte subsets identified by manual gating in healthy subjects and SLE patients before and after IL-2 immunotherapy (*n* = 12-24). **b**,**c**, Ki-67-positivity of immune cell subsets identified by manual gating during the first (b) and fourth cycle (c) of IL-2 immunotherapy. Ranked according to p-values comparing Ki-67 expression between days 0 and 5 (b) or days 63 and 68 (c) (*n* = 12-24). **d**, Ratios of frequencies and proliferation of Tfr to Tfh, Treg1 to Th1, Treg2 to Th2, and Treg17 to Th17 upon IL-2 treatment in SLE patients and untreated healthy controls (*n* = 12-24). **e**, Selected serum cytokines in healthy subjects and SLE patients before and after IL-2 treatment (*n* = 12-24). **f**, Bulk RNA sequencing of sorted Treg, CD4^+^ conventional T (Tcon), and CD8^+^ T cells depicting selected differentially expressed genes compared between days 0 and 5, and days 0 and 68. FDR < 0.05 and absolute log2 fold change > 1 and <–1 (*n* = 11-12). **g**, Enrichment plots depicting the hallmarks IL-2–STAT5 signaling, MTORC signaling, and Oxidative Phosphorylation of Treg cells between days 0 and 68 (*n* = 11-12). Shown are mean values with error bars representing the standard error of the mean (a,d,e) or median with quartiles and minimal to maximal extension (e). After normality testing, p-values were calculated with repeated-measure one-way ANOVA with Fisher’s LSD test (a,e), Friedman test with uncorrected Dunn’s test (a, d), Wilcoxon test (b, c), Mann-Whitney test (d), quasi-likelihood test on negative binomial generalized linear model (f) and permutation testing (g). Abbreviations: cMonocytes, classical monocytes; iMoncytes, intermediate monocytes; ncMonocytes, non-classical monocytes; NK, natural killer cell; RTE, recent-thymic emigrant; Tcon, conventional T cell; Tcm, central-memory T cell; Temra, CD45RA^+^ effector memory T cell; Tem, effector memory T cell; Tfh, follicular helper T cell; Tfr, follicular helper T cell; Th, helper T cell; Treg, regulatory T cell.

In recent years, progress in characterizing Treg cells has proposed populations similar to helper T cell subsets. For instance, T follicular regulatory (Tfr) cells have been shown to suppress Tfh cells, thus impairing germinal center reactions^22^. Similarly, Treg1 cells have been found to suppress T helper (Th) 1 cells^23^. Although less clear for Treg2 and Treg17 cells, it has been shown that IRF4-dependent Treg cells inhibited Th2-mediated responses^24^ and STAT3-dependent Treg cells suppressed Th17 responses^25^, with RORγt^+^ Treg cells controlling intestinal inflammation^26^. Following this notion, we assessed the expansion and proliferation of these specialized subsets, which revealed selective proliferation of Tfr over Tfh cells, Treg1 over Th1 cells, and Treg2 over Th2 cells after IL-2 treatment (**Fig. 2d**). Whereas proliferation of Treg17 was not more pronounced than that of Th17 cells upon IL-2 treatment, the ratio of circulating Treg17 over Th17 increased, possibly due to IL-2 inhibiting differentiation of Th17 cells^27^.

Measuring serum cytokines with targeted proteomics revealed an increase in immunosuppressive IL-10 and soluble programmed cell death 1 ligand 1 (PD-L1) (**Fig. 2e**), with the latter having the potential of exerting regulatory properties, as previously shown in tumors^28^. We further noted, after each of the IL-2 treatment cycles investigated, a transient increase of pro-inflammatory cytokines, including IL-6, tumor necrosis factor (TNF), and interferon gamma (IFNγ), which returned to baseline before the consecutive cycle. This was likely attributed to the significant but mild stimulation of effector T and NK cells by low-dose IL-2 immunotherapy (**Fig. 2b**,**c**) and could explain some of the observed transient adverse events, such as injection site reactions and flu-like symptoms. Conversely, IL-17A serum levels remained stable over the course of our study. Interestingly, we saw a slight decrease in IL-7, likely due to an increase of the IL-7-dependent memory T cell pool^29^. We also observed an increase in colony-stimulating factor 1 (CSF1), suggesting an indirect effect of IL-2 on monocyte proliferation (**Fig. 2c**,**e**).

To compare IL-2-induced transcriptional changes between Treg cells and the other main T cell subsets, we performed bulk RNA sequencing of fluorescence-activated cell sorting (FACS)-purified CD4^+^ CD127^lo^ CD25^+^ Treg, CD4^+^ Tcon, and CD8^+^ T cells at baseline, after the first cycle of IL-2 treatment (day 5), and at the primary endpoint (day 68) to investigate short-term and long-term effects of IL-2 immunotherapy. Comparing baseline to endpoint (day 0 vs. day 68), CD8^+^ T cells did not reveal any differentially regulated genes, and CD4^+^ Tcon cells only showed 2 down- and 24 upregulated genes (**Fig. 2f**). Of the upregulated genes, *PDCD5* is associated with Th1 and Th17 suppression, and *CD36* can promote T cell dysfunction^30,31^. In contrast, Treg cells showed marked differential gene expression after the first treatment cycle, with 8 down- and 55 upregulated genes, which became even more pronounced after 9 weeks of treatment, resulting in 8 down- and 80 upregulated genes. These included genes associated with activation and suppression (*IL2RA, HLADR, IKZF4, LRRC32*), cytokines and cytokine signaling (*SOCS1, SOCS2, IL1R1, IFNG*), proliferation (*CDKN2B*), and the TNF receptor superfamily (*TNFRSF4* [OX40], *TNFRSF8* [CD30], *TNFRSF11A* [RANK], *TNFRSF13C* [BAFFR], *TNFRSF18* [GITR]). Two of the most significantly upregulated genes included *CEACAM1*, which has been found to be upregulated by IL-2 signals^32^, and *CD300LD* (**Fig. 2f**). The latter has been described as a functional receptor for norovirus entry into mouse cells, however, its specific role in Treg cells is unclear^33^. Analysis of the hallmark gene sets in Treg cells revealed increased activation of IL-2–STAT5 signaling, MTORC signaling, and oxidative phosphorylation (**Fig. 2g**), the latter driven by FOXP3^34^. To confirm suppressive activity of expanded Treg cells, we conducted Treg cell suppression assays at baseline and primary endpoint, which showed that Treg cells retained suppressive activity despite their proliferation.

Overall, these data demonstrate that, following IL-2 immunotherapy, Treg cells proliferate vigorously and maintain their suppressive capacity. Moreover, transcriptional analysis of T cell subsets suggests that low-dose IL-2 immunotherapy primarily induces transcriptional changes in Treg cells, whereas expression profiles of CD8^+^ T and and CD4^+^ Tcon cells are not or barely affected. However, even low doses of IL-2 exert pleiotropic effects in humans, also inducing proliferation of effector-type immune cells.

### IL-2 immunotherapy preferentially expands distinct Treg cell clusters

After conducting a detailed analysis of the major immune cell subsets identified by manual gating, we switched to an exploratory approach, analyzing the Treg cell compartment by unsupervised clustering. Phenograph clustering allowed identification of all major immune cell subsets, including CD4^+^ FOXP3^+^ CD127^lo^ CD25^+^ Treg cells. Further subclustering of Treg cells identified 12 clusters with distinct phenotypes (**Fig. 3a**,**b**). A simplified trajectory extended from CD45RA^+^ Treg phenotypes on the far left to more activated CD45RA^−^ Treg phenotypes with increasing CD25 expression on the far right. Before IL-2 immunotherapy, SLE patients had less mature and less activated Treg cells compared to healthy patients. This functional deficit was corrected after IL-2 immunotherapy, with a shift toward the activated populations on the far right (**Fig. 3c**). When checking annotated clusters, we observed that RTE Treg cells (cluster 6) showed a quantitative defect compared to healthy subjects, with slow restoration during treatment (**Fig. 3d**). Cluster 11, representing CD45RA^hi^ Treg cells, increased, whereas clusters 2 and 3, comprising CD45RA^int^ Treg cells, decreased upon IL-2 immunotherapy. PD-1^hi^ Treg cells (cluster 8) were highly enriched in SLE patients and rapidly declined to physiologic levels after treatment; these cells could represent dysfunctional Treg cells, as PD-1 expression can impact their suppressive function^35^. Cluster 4, containing CD45RA^lo^ Treg cells, showed a marked reduction upon IL-2 treatment, which was likely due to their differentiation into the more activated Treg cell clusters 5 and 7. Highly differentiated Treg cells were found particularly in clusters 7 (HLA-DR^+^), 9 (CD38^+^), and 12 (CD38^+^ HLA-DR^+^), which vigorously expanded upon IL-2 immunotherapy. Interestingly, we identified two Treg cell subsets (clusters 1 and 10) that clustered closely with RTE Treg cells, which were CD31^high^, expressed low to intermediate levels of CD45RA and decreased upon IL-2 treatment. According to previous reports these cells contain few T cell receptor excision circles and are not considered RTEs^36^.

**Fig. 3.**
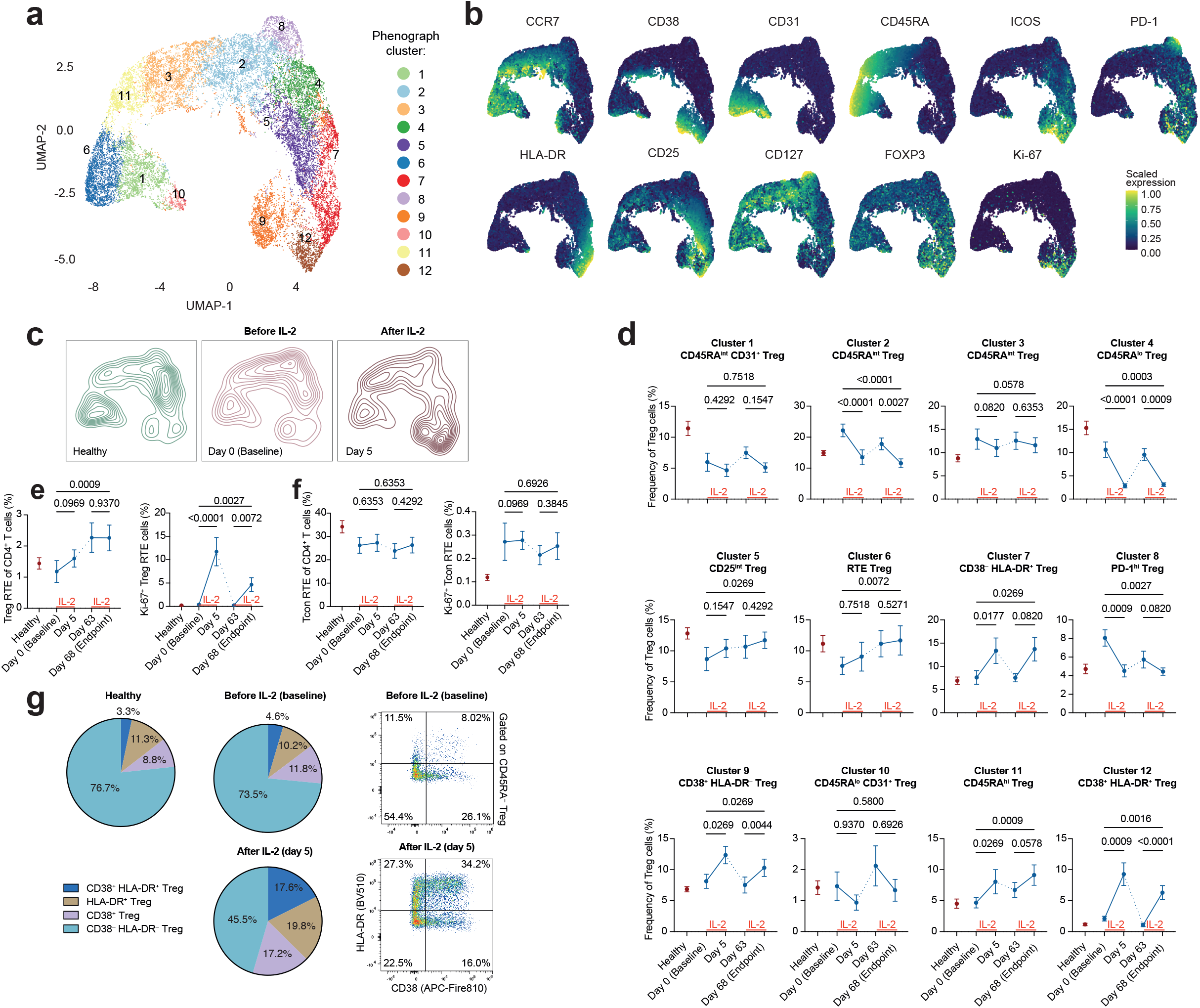
IL-2 immunotherapy preferentially expands distinct Treg cell clusters. **a**, Uniform Manifold Approximation and Projection (UMAP) representing Phenograph clusters of Treg cells of SLE patients before and after IL-2 immunotherapy and healthy controls (*n* = 12-24). **b**, Scaled expression of selected markers (*n* = 12-24). **c**, Contour plots depicting healthy subjects and SLE patients before IL-2 treatment at day 0 and after IL-2 treatment at day 5 (*n* = 12). **d**, Changes in Treg cell cluster abundancy in SLE patients before and after IL-2 immunotherapy and in healthy controls (*n* = 12-24). **e**,**f**, Frequencies and proliferation of recent-thymic emigrant (RTE) Treg (e) and RTE CD4^+^ conventional T (Tcon) cells in SLE patients before and after IL-2 treatment and in healthy controls (*n* = 12-24). **g**, Distribution of CD38^+^ HLA-DR^+^, CD38^−^ HLA-DR^+^, CD38^+^ HLA-DR^−^, and CD38^−^ HLA-DR^−^ Treg cell subsets of total Treg cells in SLE patients before and after IL-2 immunotherapy and in healthy controls (*n* = 12-24). Shown are mean values with error bars representing the standard error of the mean (d,e,f,g). After normality testing, p-values were calculated with Friedman test with uncorrected Dunn’s test (d,e,f).

As cluster 6, representing RTE Treg cells, suggested a progressive normalization of thymic Treg cell output upon IL-2 treatment, we aimed to confirm this observation by using manual gating. Indeed, CD45RA^+^ CD31^+^ RTE Treg cells were reduced in SLE patients compared to healthy individuals, suggesting a deficiency in thymic Treg cell output in SLE. IL-2 immunotherapy induced pronounced proliferation and frequencies of RTE Treg cells increased 2–3 fold (**Fig. 3e**), whereas no change was evident in CD4^+^ RTE Tcon cells (**Fig. 3f**).

Having identified clusters 7, 9, and 12 as the most IL-2-sensitive Treg cell subpopulations, we applied manual gating to identify these cells based on surface markers CD38 and HLA-DR. This allowed subdivision into four CD45RA^−^ Treg cell subsets, namely CD38^−^ HLA-DR^−^ Treg, CD38^+^ Treg, HLA-DR^+^ Treg, and CD38^+^ HLA-DR^+^ Treg cells (**Fig. 3g**). Assessment of distribution revealed that CD38^+^, HLA-DR^+^, and CD38^+^ HLA-DR^+^ Treg cells constituted approximately 25% of all CD45RA^−^ Treg cells in both healthy subjects and SLE patients at baseline, with no notable difference between these groups. Upon IL-2 treatment, these subsets significantly expanded to approximately 55%, with CD38^+^ Treg cells expanding 1.5-fold, HLA-DR^+^ Treg cells 1.9-fold, and CD38^+^ HLA-DR^+^ Treg cells even 3.8-fold.

Together, these results demonstrate differential effects of IL-2 immunotherapy on distinct Treg cell clusters. Treg cells expressing CD38 and/or HLA-DR are the most IL-2-sensitive subsets. IL-2 immunotherapy also corrects the deficiency in RTE Treg cell output in SLE.

### Single-cell RNA sequencing reveals functional properties of Treg cell subsets

To further elucidate the differential roles of Treg cell subsets, we performed single-cell RNA sequencing with cellular indexing of PBMCs in three SLE patients before and after IL-2 treatment (day 0 and day 5). Primary FlowSOM clustering based on surface protein expression allowed identification of all major immune cell subsets, including Treg cells.

Secondary weighted nearest neighbor clustering of protein markers of Treg cells revealed clusters reflecting the CD45RA^+^ Treg, CD45RA^−^ CD38^−^ HLA-DR^−^ Treg, CD38^+^ Treg, HLA-DR^+^ Treg, and CD38^+^ HLA-DR^+^ Treg cell subsets, which matched the subsets we previously identified by flow cytometry (**Fig. 4a**,**b**). Analysis of RNA expression distribution between the CD38^+^ Treg, HLA-DR^+^ Treg, and CD38^+^ HLA-DR^+^ Treg cell subsets demonstrated that the majority of genes were expressed by all three subsets (n = 3,325), while 1,664 genes were uniquely expressed by only one of the subsets (**Fig. 4c**). We performed differential analysis of protein and RNA expression between the three subsets. Hierarchical clustering of all significant surface proteins (**Fig. 4d**) and the top 50 genes (**Fig. 4e**) showed distinct activation programs by the three subsets. These included proteins and genes associated with migration (CD62L, ITGB1, ITGA6, ITGB7, CCR4, CCR6, CXCR3, *CCR9*) and with activation and suppression (ICOS, CD25, CD39) (**Fig. 4d**,**e**).

**Fig. 4.**
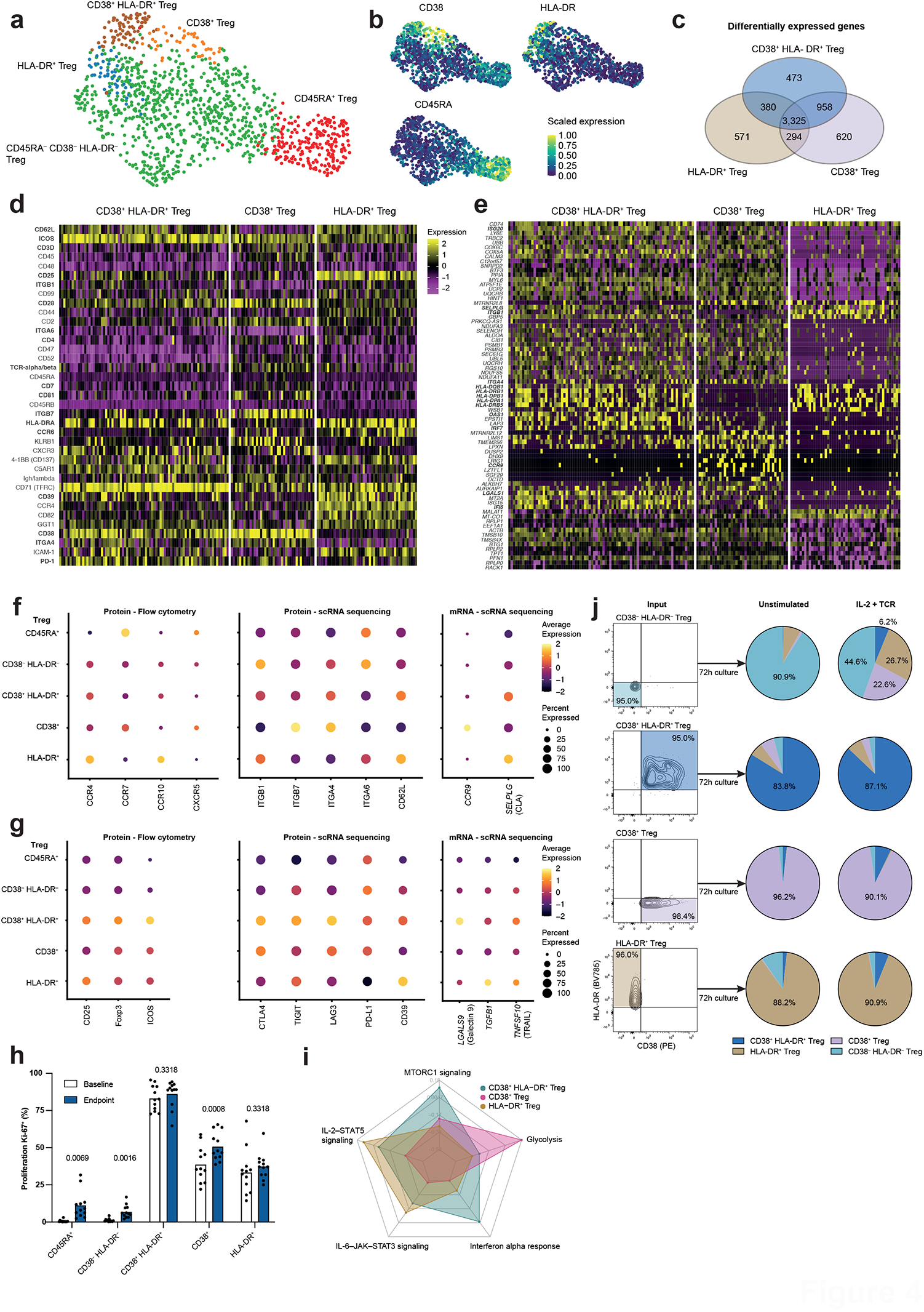
Single-cell RNA sequencing reveals functional properties of Treg cell subsets. **a**,**b**, Weighted nearest neighbor clustering of Treg cells from three SLE patients before and after IL-2 immunotherapy (a) and with scaled marker expression (b) (*n* = 3). **c**, Counts of differentially expressed genes comparing CD38^+^ HLA-DR^+^, CD38^+^, and HLA-DR^+^ Treg cell subsets (*n* = 3). **d**,**e**, Heatmaps depicting significantly differentially regulated surface proteins (d) and top 50 differentially expressed genes (e) of CD38^+^ HLA-DR^+^, CD38^+^, and HLA-DR^+^ Treg cells (*n* = 3). **f**,**g**, Dot plots depicting surface protein expression measured by flow cytometry (left panel, *n* = 12, days 0 and 5), protein expression quantified by single-cell RNA sequencing (*n* = 3, days 0 and 5), and gene expression quantified by single-cell RNA sequencing (*n* = 3, days 0 and 5) for proteins and genes associated with migration (f) and activation and suppression (g) in indicated Treg cell subsets. **h**, Proliferation of Treg cell subsets measured by Ki-67 expression before and after IL-2 immunotherapy (*n* = 12). **i**, Normalized expression of hallmarks MTORC1 signaling, glycolysis, interferon alpha response, IL-6–JAK–STAT3 signaling, and IL-2–STAT5 signaling of indicated Treg cell subsets (*n* = 3). **j**, *In vitro* differentiation of FACS-sorted CD38^+^ HLA-DR^+^, CD38^+^, and HLA-DR^+^ Treg cell subsets cultured for 72 hours either in control medium or in presence of IL-2 (500 IU/ml) and T cell receptor stimulation (anti-CD3 and anti-CD28 antibodies) (*n* = 4). Shown are mean values (h,j). After normality testing, p-values were calculated with multiple paired t-tests with Holm-Sidak test (h).

To gain detailed insight into the distinct activation states of the CD38^+^ Treg, HLA-DR^+^ Treg, and CD38^+^ HLA-DR^+^ Treg cell subsets, we compared surface protein expression, single-cell RNA sequencing protein abundance, and single-cell RNA sequencing mRNA expression of markers associated with migration (**Fig. 4f**) and with activation and suppression (**Fig. 4g**). These revealed that CD38^+^ Treg cells expressed high levels of ITGA4 and ITGB7 together with *CCR9* (**Fig. 4f**), which are implicated in gut homing^37,38^. A fraction of these cells also expressed high CXCR5 and CCR7 (**Fig. 4f**), which indicated an enrichment of Tfr cells in this subset. CD38^+^ Treg cells also carried proteins associated with suppression, including CTLA-4, LAG3, and PD-L1 (**Fig. 4g**)^39,40^. Differently, HLA-DR^+^ Treg cells displayed a skin-homing profile, with increased expression of CCR4, CCR10, and *SELPLG* (coding for CLA), as well as ITGA6 and ITGB1 (binding laminin)^37,38^. A part of this subset further expressed CD62L, indicating homing to secondary lymphoid organs (**Fig. 4f**). HLA-DR^+^ Treg cells expressed increased CD39 and *TGFB1*, thus indicating a distinct suppressive profile different from the other two subsets (**Fig. 4g**). CD38^+^ HLA-DR^+^ Treg cells did not show a discernible tissue-homing profile (**Fig. 4f**), but they carried the highest expression levels of Treg cell activation-associated markers CD25, FOXP3, ICOS, CTLA-4, TIGIT, LAG3, *LAGLS9* (coding for Galectin 9), and *TNFRSF9* (coding for TRAIL) (**Fig. 4g**), most of which have been associated with suppression^40^. This subset was the most proliferative Treg cell subset during low-dose IL-2 immunotherapy, compared to CD38^+^ and HLA-DR^+^ Treg cells. Interestingly, proliferation was absent in CD45RA^−^ CD38^−^ HLA-DR^−^ and CD45RA^+^ Treg cells (**Fig. 4h**).

Gene set variance analysis displaying hallmark gene sets on a radar chart revealed enrichment of the glycolysis gene set in CD38^+^ Treg cells (**Fig. 4i**). The IL-2–STAT5 and IL-6–JAK–STAT3 signaling gene sets were enriched in HLA-DR^+^ Treg cells, whereas MTORC1 and interferon alpha gene sets were enriched in CD38^+^ HLA-DR^+^ Treg cells. This further supports distinct activation programs induced by different cytokine signals.

To further investigate the plasticity of these Treg cell subsets, we FACS-purified CD45RA^−^ CD38^−^ HLA-DR^−^ Treg, CD38^+^ Treg, HLA-DR^+^ Treg, and CD38^+^ HLA-DR^+^ Treg subsets. Upon *in vitro* stimulation by IL-2, T cell receptor (TCR) engagement or IL-2 and TCR engagement for 72 hours, the CD38^+^ Treg, HLA-DR^+^ Treg, and CD38^+^ HLA-DR^+^ Treg subsets maintained their respective phenotypes. In contrast, CD45RA^−^ CD38^−^ HLA-DR^−^ Treg cells upregulated HLA-DR upon IL-2 stimulation, and CD38 upon combined IL-2 and TCR stimulation. These data suggested that CD38 and HLA-DR identified stable Treg cell subsets, whereas HLA-DR was induced by IL-2 signals and CD38 by combined IL-2R and TCR engagement (**Fig. 4j**).

Taken together, these results support IL-2-driven Treg cell subsets with distinct transcriptional programs of tissue homing and suppression. Whereas CD38^+^ Treg cells carry features of gut-homing cells and HLA-DR marks skin-homing Treg cells, CD38^+^ HLA-DR^+^ Treg cells constitute a highly activated and proliferative subset.

## DISCUSSION

Although SLE is a serious autoimmune disease impacting the quality of life of women of childbearing age and associated with high socioeconomic costs, only two novel drugs for the treatment of SLE have been approved in the past 60 years^13^. Belimumab is a monoclonal antibody targeting B cell-activating factor (BAFF) that reduces plasma cell differentiation and survival, while anifrolumab targets the type-I interferon (IFN) pathway activated in SLE^41,42^. Despite these recent approvals, many patients insufficiently respond to current therapies, which is likely due to the heterogeneity of SLE. One interesting and complementary approach aims to correct the functional Treg cell deficiency observed in SLE by low-dose IL-2 immunotherapy. Our data confirm previous reports of pronounced Treg cell expansion by IL-2 immunotherapy in SLE, coinciding with clinical response measured with validated clinical scores and improvement of serum biomarkers. We further demonstrated – for the first time – that low-dose IL-2 immunotherapy expanded Treg cells in a peripheral tissue, namely the skin. Interestingly, this effect appeared to be selective for Treg cells and did not affect CD4^+^ Tcon or CD8^+^ T cells. Mechanistically, IL-2 immunotherapy did not induce significant proliferation of skin-resident Treg cells; rather, it induced proliferation of CLA-expressing skin-homing Treg cells, which subsequently infiltrated the skin. This was further supported by the close interaction of Treg cells with vascular endothelial cells upon IL-2 treatment, and this relationship, in turn, might position Treg cells as gatekeepers of effector T cells trying to enter the skin. It is interesting to speculate that IL-2-stimulated skin-homing Treg cells might become independent of IL-2 upon skin infiltration by relying on signals by IL-7 and other cytokines^29,43^. This might allow persistence and long-lasting activity of skin-homing and possibly also other tissular Treg cells even after discontinuing IL-2 treatment.

To our knowledge, the current study provides the most comprehensive immune cell phenotyping to date of patients treated with low-dose IL-2 immunotherapy. Surprisingly, our data revealed that not Treg cells but CD56^bright^ NK cells were the most IL-2-sensitive immune cells, showing up to 60% proliferation after IL-2 treatment. NK cells have been reported to exert regulatory functions in multiple sclerosis patients treated with the anti-CD25 antibody daclizumab, potentially via direct cytotoxicity on autoreactive CD4^+^ T cells^44,45^. To which extent these functions also operate in SLE patients treated by IL-2 remains unclear. Although to a lesser degree, also CD4^+^ and CD8^+^ T cell subsets proliferated upon IL-2 immunotherapy. Most commonly, increased cell proliferation does not coincide with expansion in the blood, likely due to migration to secondary lymphoid organs or into the tissue. This supports our notion that human studies investigating the effects of specific immunomodulatory treatments, including stimulatory cytokines, anti-cytokine antibodies, and immune checkpoint inhibitors, should, in addition to reporting frequencies, also focus on proliferation. Furthermore, our results demonstrated that IL-2 treatment increased blood levels of both suppressive cytokines, such as IL-10, and of pro-inflammatory cytokines, such as IL-6, TNF, and IFNγ. The increase in IFNγ could be attributed to the significant expansion of CD56^bright^ NK cells^45^. However, this effect was mild and transient.

Detailed and unsupervised analysis of Treg cell phenotypes showed a strong IL-2-driven expansion of CD38^+^, HLA-DR^+^, and CD38^+^ HLA-DR^+^ Treg cells. Interestingly, proliferation during steady state was limited to these three CD45RA^−^ subsets, whereas CD45RA^+^ and CD45RA^−^ CD38^−^ HLA-DR^−^ did not proliferate. Extended phenotyping with single-cell RNA sequencing revealed these subsets showed characteristics of suppressive function and migration to specific tissues. While CD38^+^ Treg cells expressed receptors associated with gut homing, HLA-DR^+^ Treg cells expressed skin-homing receptors. On the other hand, concomitant CD38 and HLA-DR positivity marked highly proliferative Treg cells. The identification of these subsets offers a clear distinction between resting and highly activated Treg cells, the latter being fueled by IL-2 immunotherapy. Previous results in humans suggested that HLA-DR^+^ Treg cells were highly suppressive^46^, whereas CD38 correlated with a suppressive Treg cell phenotype in lupus-prone mice^47^. A detailed description of CD38^+^ HLA-DR^+^ Treg cells and their interrelation with CD38^+^ and HLA-DR^+^ Treg cells remains to be studied. Future studies should also investigate whether these subsets are impaired in different autoimmune diseases or enriched in tumors with poor outcome or resistance to immunotherapy. A first step in this direction comes from a recent study that associated poor prognosis in colorectal cancer patients with tumor-infiltrating CD38^+^ Treg cells^48^. This finding further supports the gut-homing properties of these cells. Moreover, the identification of these cells has specific implications for novel immunotherapeutic strategies adopted from oncology. In particular, the potential benefit of depleting CD38^+^ long-lived plasma cells with the CD38-targeting antibody daratumumab in SLE might be compromised by simultaneous depletion of activated CD38^+^ Treg cells^49,50^.

From a clinical perspective, one limitation of this study is the lack of a control group, thus preventing clear conclusions on the clinical response in these patients. However, the study did not aim at showing clinical efficacy and the clean study design, with blood draws before and after a 5-day course of IL-2 treatment, allows comprehensive investigation of IL-2-induced biological changes. The study also revealed limitations of low-dose IL-2 treatment. Due to the imperfect bias toward the trimeric IL-2R, stimulation of effector cell populations including NK and T cell subsets could limit its efficacy. Despite these effects, Treg cell expansion seems to be beneficial, which supports the clinical development of improved IL-2 formulations with CD25 bias^51,52^. Furthermore, the implication and mechanism of the observed B cell proliferation remains unclear. Future in-depth studies analyzing the B cell compartment upon IL-2 immunotherapy might reveal novel aspects of direct and indirect effects of IL-2 on subsets of this compartment. Of particular interest could be the subset of regulatory B cells, which has been proposed to express functional IL-2R^53^.

In summary, this constitutes the first study aiming at comprehensively phenotyping IL-2-induced effects of low-dose IL-2 immunotherapy. Categorizing Treg cells based on expression patterns of CD45RA, CD38, and HLA-DR has distinct advantages compared to previous approaches that focused on gradual expression patterns of CD25 and FOXP3. Our findings thus have important implications for the future development of improved IL-2 treatments and the characterization of Treg cells in autoimmune diseases and cancer.

## METHODS

### Study design

Within the trial “Open-label, Monocentric, Phase II, Investigator-initiated Clinical Trial on Unbiased Characterization of Immunological Parameters in Interleukin-2-treated Systemic Lupus Erythematosus” (Charact-IL-2) 12 female SLE patients were screened for inclusion and exclusion criteria and subsequently enrolled into the clinical trial. Detailed inclusion and exclusion criteria are published on ClinicalTrials.gov, accessible under the identifier NCT03312335. Briefly summarized, eligible adult participants required oral and written informed consent before enrollment into the trial, a confirmed diagnosis of SLE according to the American College of Rheumatology classification criteria^54^, stable corticosteroid dose for at least 4 weeks prior to enrollment, unchanged immunosuppressive medication for at least 4 weeks prior to enrollment, no major impairment in cardiac, pulmonary, renal, and hepatic organ functions. Exclusion criteria involved contraindication to treatment with IL-2, solid organ transplant recipient, exposure to rituximab or cyclophosphamide within 3 months before enrollment, concomitant medications above the indicated maximal dose (hydroxychloroquine >400 mg/day, prednisone >20 mg/day, azathioprine >2.5 mg/kg/day, mycophenolic acid >3 g/day, methotrexate >20 mg subcutaneously applied once weekly, belimumab after induction >10 mg/kg every 4 weeks, and simultaneous use of sirolimus and tacrolimus), history of thrombotic thrombocytopenic purpura, hemolytic-uremic syndrome or thrombotic microangiopathy, any active uncontrolled infection, pregnant or breast feeding women, lack of safe contraception, other clinically significant concomitant disease states (e.g. renal failure, hepatic dysfunction, cardiovascular disease), and known or suspected non-compliance.

Within this single-armed trial patients were treated with 4 cycles of 5 consecutive daily injections of 1.5 million international units of the human recombinant IL-2 Proleukin® (Aldesleukin, Novartis Pharma) over a time period of 68 days (9 weeks), followed by follow-up visits at days 96 and 124. Treatment cycles were initiated at days 0, 21, 42, and 63 with a maximal accepted tolerance of ±3 days. The first injection was done at the Department of Immunology, University Hospital Zurich with the 4 remaining injections done independently by the patients at home. Study visits including medical history assessment, clinical examinations, adverse events recording, study drug log, vital signs, blood draws, and urine samples for routine and experimental laboratory assessments were conducted during eligibility screening (up to 8 weeks before treatment initiation), at the beginning of each treatment cycle (days 0, 21, 42, and 63) and one day after the last injection of IL-2 during each treatment cycle (days 5, 26, 47, and 68), as well during follow-up visits (days 96 and 124). Optional skin punch biopsies (5 mm diameter) were obtained from 4 patients from non-lesional skin at days 0 and 68, with the second sample taken with an approximate distance of 3 cm from the first sample.

The primary outcome of the study was an increase in percentage of Treg cells of total CD4^+^ T cells between day 0 (baseline) and day 68 (visit 9) measured in the blood of SLE patients not receiving belimumab treatment, tested using a paired t-test. Treg cells were defined as CD3+ CD4+ CD127^lo^ CD25^hi^ Foxp3^+^ and total CD4^+^ T cells are defined as CD3^+^ CD4^+^. Secondary outcomes included an exploratory assessment of cellular and soluble factors in the blood before and after IL-2 immunotherapy, assessment of immune cells in skin punch biopsies collected at days 0 and 68, clinical response measured with SELENA-SLEDAI, BILAG-2004 and Physician’s Global Assessment scores, as well the calculated SLE Responder Index (SRI) at days 0, 26, 68, and 124^55-58^, dose of concomitant medication, and safety with adverse events recording according to the Common Terminology Criteria for Adverse Events version 5.0.

The sample size was calculated assuming an expected increase of Treg cells by 8% (standard deviation = 5.8%) between baseline and visit 9^11^. With a level of significance α = 0.05 and 90 % power a sample size of N = 8 was calculated. With an expected maximal dropout rate of one third we defined 12 SLE patients to be recruited into the clinical trial to meet the primary endpoint.

The study was carried out in accordance with principles enunciated in the current version of the Declaration of Helsinki, the guidelines of Good Clinical Practice issued by International Council for Harmonisation of Technical Requirements for Pharmaceuticals for Human Use, Swiss legal and Swiss competent authority’s requirements. Before initiation, the trial protocol was approved by the ethical committee of the Canton of Zurich, Switzerland (Swiss National Clinical Trials Portal registration number SNCTP000002372) and the Swiss Agency for Therapeutic Products Swissmedic. After enrollment of the first patient into the trial there were no changes to the methods or trial outcome, or any other changes to the protocol. Data was exclusively collected at the Department of Immunology, University Hospital Zurich with key clinical parameters including primary endpoint outcome entered into a validate electronic case report file (secuTrial® 5.4.3.5, interactive Systems GmbH). For assurance of compliance with legal requirements and quality assurance the trial was independently monitored by the Clinical Trials Center of the University Hospital Zurich.

### Human samples

Human samples were collected within the clinical trial and for healthy controls withing the “Fundamental research project for phenotypical and functional characterization of different leukocyte subsets in healthy and diseased individuals” (approved by the ethics committee of the Canton of Zurich, BASEC registration number 2016-01440) or obtained from the blood donation network Basel, Switzerland (Blutspende SRK Beider Basel). Prior to sample collection, written informed consent was obtained. Human blood was collected into EDTA Vacutainer tubes (BD Biosciences) followed by Ficoll-Paque PLUS (GE Healthcare) gradient centrifugation for peripheral blood mononuclear cell (PBMC) isolation. Isolated PBMCs were frozen in fetal calf serum (FCS, Gibco) containing 10% dimethyl sulfoxide (Sigma) and stored for less than 3 years in liquid nitrogen prior to analysis. Serum was isolated from blood collected with Clot Activator Vacutainer tubes (BD Biosciences) and stored for less than 3 years at –80°C prior to analysis.

Skin punch biopsies were taken according to standard procedure at the Department of Immunology, University Hospital Zurich, embedded in Optimal Cutting Temperature Compound (Tissue-Tek) and snap-frozen in liquid nitrogen (time from biopsy to freezing approximately 30 min, stored for less than 2 years at –80 °C before analysis).

### Flow cytometry

Frozen PBMC samples were processed according to standard protocols. In brief, after thawing in pre-warmed complete medium containing 10% FCS, the cells were plated in 96-well plates. For the primary endpoint staining cells were initially stained with Fixable Viability Stain diluted in phosphate-buffered saline (PBS) at 4 °C for 30min, subsequently washed with FACS buffer (PBS with 2% FCS, 2 mM EDTA) and stained with surface staining for 30 min at 4 °C. Cells were fixed and permeabilized using the Foxp3 / Transcription Factor Staining Buffer Set (eBioscience, ThermoFisher) for 30 min at 4 °C, before intracellular staining in PermWash solution for 30 min at room temperature. For the immunophenotyping panel (Figures 2 and 3), after Live-Dead cell staining and Fc block, cells were stained with the chemokine receptor staining mix for 20 min at 37°C, followed by the remaining surface staining for 30 min at room temperature, followed by fixation and permeabilization and intracellular staining. Staining mixes were centrifuged at 14,000 g for 2 min before staining to remove aggregates.

Samples were acquired with a BD LSRFortessa using BD FACSDiva software (BD Biosciences) for the primary endpoint staining, or Cytek Aurora using SpectroFlo software (Cytek Biosciences) for the immunophenotyping panel. Quality control for the spectral flow cytometer was performed daily. All longitudinal samples were acquired in the same batch, additionally in each batch the same healthy control sample was recorded to ensure consistent results.

### Flow cytometry data analysis

The flow cytometry data was analyzed using FlowJo software (version 10.8.0) for manual gating and R (version 4.1.2) for unsupervised analysis. Unsupervised analysis was performed following the CATALYST workflow (CATALYST package, version 1.18.1). In brief, Tregs were identified using primary and secondary FlowSOM clustering (FlowSOM package, version 2.2.0). Tregs were further subclustered using Phenograph clustering (Rphenograph package, version 0.99.1).

### Imaging mass cytometry

Histological skin sections from punch biopsies were processed for staining with a 38-marker panel as previously described^59,60^. Where available, pre-conjugated antibodies were used with the remaining antibodies conjugated in-house with heavy metal isotopes using the Maxpar Antibody Labeling Kit following standard protocols (Fluidigm). The conjugation of antibodies against α-smooth muscle actin (SMA) and collagen I with cisplatin 194 and 198 were performed following an adapted published protocol^61^. Tissue sections with 5-μm thickness were dried at room temperature, followed by fixation with 1% paraformaldehyde for 5 min at room temperature. After cooling in methanol for 5 min at –20 °C, sections were incubated with the antibody mixture overnight at 4°C. The following day, sections were washed three times with staining buffer for 5 min and once with deionized water for 2 min, and subsequently air dried. Skin sections were then ablated at 200 Hz on a Helios time-of-flight mass cytometer coupled to a Hyperion Imaging System (Fluidigm), performed according to standard operating procedures by Fluidigm.

Single-cell segmentation was done following the Imc Segmentation Pipeline (version 2.3). Briefly, Ilastik (version 1.3.3post3) was used for pixel classification and creation of probability maps. These were then segmented into single cells using CellProfiler (version 4.2.1). An additional step of histogram equalization (256 bins and kernel size 17) was added as the first step of image segmentation. Nuclei were segmented by two class Otsu thresholding with a thresholding correction factor of 1.7. Subsequently, cell masks were generated by expanding the nuclear masks by three pixels. The CellProfiler module “measure neighbors” was used to define cells with a maximum distance of 8 µm between cell boarders of neighboring cells.

All downstream analysis was conducted in R (version 4.1.2). FlowSOM clustering was used for clustering-based identification of main cell lineages as implemented by the CATALYST package (version 1.18.1). The secondary clustering was performed with the Rphenograph package (version 0.99.1). Visualization of the IMC data were created with the cytomapper package (version 1.6.0). Interaction maps were created using the package imcRtools (version 1.0.2). ImcRtools was used to quantify the interactions using method “classic”.

### Treg suppression assay

CD4^+^ CD127^−^ CD25^+^ Treg and CD4^+^ CD127^−/+^ CD25^−^ Tcon cells from patients at day 0 (baseline) and day 68 (endpoint) were FACS-sorted and labeled with CellTrace violet and CellTrace CFSE (ThermoFisher), respectively. Cells were then incubated at defined ratios in U-bottom plates coated with anti-CD3 (OKT3, Biolegend, 5µg/ml) for 5 days in complete RPMI medium without IL-2. CellTrace dilution was measured with flow cytometry and proliferation indices calculated.

### Treg differentiation assay

FACS-sorted CD45RA^−^ CD38^−^ HLA-DR^−^ Treg, CD38^+^ Treg, HLA-DR^+^ Treg, and CD38^+^ HLA-DR^+^ Treg cell subsets were incubated in U-bottom plates containing complete RPMI medium for 3 days. Conditions included medium only (control), IL-2 (aldesleukin, Novartis Pharma, 500 IU/ml), TCR stimulation (plate precoated with anti-CD3 [OKT3, Biolegend, 10mg/ml] and anti-CD28 [CD28.2, Biolegend, 1mg/ml]), or IL-2 combined with TCR stimulation.

### Olink cytokine measurements

Human serum samples before and after IL-2 treatment were collected and stored as described before. Human cytokines were measured at the Swiss Institute of Allergy and Asthma Research with a proximity extension assay using the Olink Inflammation Panel according to standard protocol (Olink).

### Bulk RNA sequencing

PBMCs were sorted using a BD FACS Aria III into three fractions: CD3^+^ CD4^+^ CD8^−^ CD127^low^ CD25^+^ Treg cells, CD3^+^ CD4^+^ CD8^−^ CD127^+^ CD25^−^ Tcon cells, and CD3^+^ CD4^−^ CD8^+^ T cells. The cells were sorted directly into RLT Plus lysis buffer (Qiagen) containing 1% 2-mercaptoethanol (Sigma-Aldrich), followed by storing at -80°C. The RNeasy Plus Micro Kit (QIAGEN) was used according to manufacturer’s instruction to isolate RNA. RNA Sequencing and data preprocessing was performed by the Functional Genomic Center Zurich. Library preparation was done using the SmartSeq2-Picelli method. Samples were sequenced on an Illumina NovaSeq 6000 System. The resulting raw FASTQ files were aligned to GRCh38.p13 reference transcriptome with gene model from GENCODE (release 32) using Kallisto sequence aligner (version 0.46.1).

All downstream analyses were conducted in R (version 4.1.2). Differential gene expression was performed with the package edgeR (version 3.36.0). First, Gene libraries were filtered and normalized as described in the edgeR vignette and the F statistic was calculated using edgeR’s glmQLFTest function with a design matrix including time point and patient ID and adjusted for false discovery rate. The F statistic from the differential gene expression was multiplied with the sign of the logFC to create a pre-ranked gene list. Gene Set Enrichment Analysis was performed on this pre-ranked gene list using the R package FGSEA (version 1.20.0). We used the FGSEA-multilevel procedure with 100’000 permutations and the Hallmark gene set from the Molecular Signature Databases available in R through the msigdbr package (version 7.4.1) and set a seed of 42 (set.seed(42) in R) for reproducibility. The resulting p values were corrected for multiple testing with the Benjamini-Hochberg procedure.

### Single-cell RNA sequencing

Single-cell RNA sequencing was done as previously described^62^. Frozen PBMCs were thawed as described above and 400,000 live cells plated in a 96-well plate. Cells were washed, resuspended in 22.5 μl FACS buffer and incubated with 2.5 μl of TruStain FcX (Biolegend) for 10 min at 4 °C. Subsequently, 22.5 μl TotalSeq-C Human Universal Cocktail, V1.0 (Biolegend) supplemented with additional TotalSeq-C antibodies, including a hashtag for sample multiplexing, and the staining antibodies was added and incubated for 30 min at 4 °C. Prior to staining, the TotalSeq-C Human Universal Cocktail was reconstituted, and the antibody mix centrifuged at 14,000 g for 10 min according to manufacturer’s instruction. Afterwards, cells were washed three times with 1.4 ml FACS buffer, then resuspended and FACS-sorted (live, CD45^+^ non-granulocytes) on a FACS Aria III 4L sorter using an 85 μm nozzle. The cells were sorted into separate tubes, which after sorting were pelleted at 300 g for 10 min at 4 °C. The cells were processed for single-cell RNA sequencing using the 5′ Single Cell GEX and VDJ v1.1 platform (10x Genomics). For this, the cells were resuspended in loading buffer (PBS with 0.04% BSA), counted, mixed in equal numbers, and subsequently loaded into the 10x Chromium Chip following manufacturer’s instructions. 13 cycles of initial cDNA amplification were performed and individual libraries for gene expression (GEX), B cell receptor (BCR), T cell receptor (TCR) and surface protein targeting antibody derived tags (ADTs) were created. Finally, libraries were quantified using a Qubit Fluorometer, pooled in a ratio of 6:1:1:1 (GEX:VDJ-BCR:VDJ-TCR:ADT) and sequenced on a NovaSeq 6000 system. Demultiplexed FASTQ files were preprocessed using 10x Genomics’ “cellranger multi” pipeline (alignment to human reference genome GRCh38) v5.0 and parameter “force-cells” set to 20,000.

All downstream analysis was conducted in R (version 4.1.2). Quality control, demultiplexing and data integration followed the workflow described by the Seurat package (version 4.1.1)^63^. Briefly, only cells with a feature count between 400 and 2,500 and mitochondrial content of less than 10% were retained. After library size normalization, sample demultiplexing was done using Seurat’s HTODemux function. As an additional quality control to exclude low quality cells and cells with unknown sample ID, we used the souporcell package (v.2.0) and its internal pipeline to cluster single cell transcriptomes by genotypes and matched resulting souporcell cluster IDs to HTO derived patient IDs. Following this, the three runs were integrated by selecting variable features as integration anchors.

For clustering-based cell type identification FlowSOM (FlowSOM package, version 1.4.0) and Consensus Clustering (ConsensusClusterPlus package, version 1.60.0) were used to identify main cell lineages prior to weighted nearest-neighbor clustering (Seurat package) to identify subclusters. DGE was calculated as implemented by Seurat with the FindMarkers function. Gene Set Enrichment Analysis was performed on these DGE results using the package FGSEA as described for bulk RNA sequencing.

### Software

Specific software used for analysis is mentioned where applied. One cartoon was created using Biorender.com.

### Statistical analyses

Detailed information on sample numbers and statistical tests used are described in the figure legends. Calculation was performed with the GraphPad Prism 9.4.1 or R 4.1.2 and all tests were two-sided. Individual packages are specified where used. Individual packages are specified where they were used. In general, differences between two groups were tested with paired t-tests for normally distributed data or Wilcoxon test for not normally distributed data. Multiple comparison was done with either one-way ANOVA with Fishers LSD test for normally distributed data or Friedman test with uncorrected Dunn’s test for not normally distributed data. Normality testing was done using d’Agostino-Pearson test. For all statistical analyses, significance was accepted at the 95% confidence level (P < 0.05). Exact *P*-values are provided.

## Data Availability

The raw datasets generated by imaging mass cytometry as well as bulk and single-cell RNA sequencing will be deposited upon publication in a peer reviewed journal. All other data and materials are available upon request from the corresponding author after publication of this study in a peer reviewed journal.

## ACKNOWLEDGMENTS

We thank our patients for participating in this trial. We thank Laura Bürgi for help with experiments, Elisabeth Käser and Corina Defila for their support with patient care, Daniela Impellizzieri, Vincent van Unen and the members of the Boyman laboratory for helpful discussions and support with experimental designs.

## AUTHOR CONTRIBUTIONS

M.R. conceived the project, conducted the clinical trial, collected data, designed and performed experiments, and analyzed and interpreted data. Y.Z. performed experiments, analyzed data. D.C. analyzed data and performed experiments. N.G. performed imaging mass cytometry experiments. J.S. collected and analyzed data. J.M. performed single-cell RNA sequencing experiments. U.S. supported the clinical trial and patient recruitment. A.M. designed single-cell RNA sequencing experiments. F.K. designed imaging mass cytometry experiments. O.B. conceived the project, designed experiments, and interpreted data. M.R. and O.B. wrote the manuscript with contributions from all the authors. The final draft of the article was approved by all the authors.

